# Ethambutol resistance preceding macrolide resistance in *Mycobacterium avium* complex pulmonary disease: a retrospective longitudinal study and in vitro analysis

**DOI:** 10.64898/2026.08.12.26360314

**Authors:** Masashi Ito, Fumiya Watanabe, Asami Osugi, Akio Aono, Keiji Fujiwara, Koji Furuuchi, Tatsuya Kodama, Takashi Ohe, Takashi Yoshiyama, Shoji Kudoh, Satoshi Mitarai, Kozo Morimoto

## Abstract

**Objectives:** To investigate whether ethambutol resistance in *Mycobacterium avium* complex is associated with the emergence of macrolide resistance.

**Methods:** Patients who developed macrolide resistance during guideline-based treatment were included, and longitudinal analyses of minimum inhibitory concentrations and mutations in *embB* or the upstream region of *embA* were performed. Clinical, microbiological, and radiological characteristics were compared according to the mutation status of *embB* or *embA* upstream region, prior to the emergence of macrolide resistance. We further evaluated the impact of *embB* mutation on the development of macrolide resistance using *in vitro* time-kill assays.

**Results:** Sixteen patients developed macrolide resistance during guideline-based treatment. None of these patients had an ethambutol minimum inhibitory concentration ≥16 μg/mL or *embB* or *embA* upstream mutations at treatment initiation; however, 8/16 patients (50.0%) had an ethambutol minimum inhibitory concentration ≥16 μg/mL at the time of macrolide resistance detection, and 7/16 (43.8%) had developed *embB* or *embA* upstream mutations prior to the emergence of macrolide resistance. Cavitary lesions were present in 1/7 (14.3%) patients with *embB* or *embA* upstream mutations. In strains with *embB* mutations, the minimum inhibitory concentration of ethambutol increased by 1–2 dilutions relative to that of pretreatment isolates, with a corresponding increase in the concentration required to suppress macrolide resistance.

**Conclusions:** Ethambutol resistance may contribute to the development of macrolide resistance in patients with *M. avium* complex pulmonary disease, particularly in those without cavitary lesions.

## Introduction

The incidence and prevalence of nontuberculous mycobacterial pulmonary disease (NTM-PD) are increasing worldwide. Globally, the most prevalent nontuberculous mycobacteria (NTM) are *M. avium* and *M. intracellulare*, which are collectively referred to as *Mycobacterium avium* complex (MAC) [1]. In both the international and British Thoracic Society guidelines, a three-drug regimen including a macrolide is recommended for the treatment of MAC pulmonary disease (MAC-PD) [2, 3]. The treatment success rate for guideline-based treatment was reported to be 65.7% in macrolide-susceptible patients with previously untreated MAC-PD [4]. In contrast, treatment outcomes for macrolide-resistant MAC-PD are poor, with sputum culture conversion rates of only 11–15% [5, 6].

It has been reported that 22% of patients with refractory MAC-PD develop macrolide resistance [7]. The use of treatment regimens without ethambutol has been identified as a risk factor for the development of macrolide resistance [6, 8]. However, even guideline-based treatment that includes ethambutol can lead to macrolide resistance in 9–12% of patients with MAC-PD [6, 9–11]. The mechanisms underlying macrolide resistance during guideline-based treatment remain unclear.

A recent report revealed cases in which the minimum inhibitory concentration (MIC) of ethambutol increased during treatment for MAC-PD, leading to the development of ethambutol resistance [12]. Mutations in *embB* and the upstream region of *embA* have been suggested as contributors to ethambutol resistance in *Mycobacterium tuberculosis*, as well as MAC-PD [14]. However, the frequency and distribution of *embB* or *embA* upstream mutations, as well as the clinically relevant cutoff value of the ethambutol MIC in patients with MAC-PD, have not yet been clearly established. Furthermore, the effects of ethambutol resistance on treatment outcomes in patients with MAC-PD remain controversial [15–17].

We hypothesized that ethambutol resistance may precede macrolide resistance.We longitudinally analysed ethambutol MICs and the emergence of embB or embA upstream mutations in patients who developed macrolide resistance during guideline-based treatment. Using a recently reported in vitro time-kill assay [13], we experimentally assessed the effect of *embB* mutations on the ability of ethambutol to suppress macrolide resistance in clinical isolates.

## Methods

### Study design and patients

This study included patients with MAC-PD who underwent antimicrobial susceptibility testing using BrothMIC SGM^®^ (Kyokuto Pharmaceutical Industrial Co., Ltd., Tokyo, Japan) at Fukujuji Hospital between October 2022 and February 2024. MIC data for clarithromycin, ethambutol, and rifampicin were collected.

Of the 345 patients with MAC-PD who underwent antimicrobial susceptibility testing, 23 newly developed macrolide resistance during treatment. We excluded five patients who had received inappropriate treatment without ethambutol for more than 1 month before the development of macrolide resistance, as well as two patients for whom isolates obtained before the emergence of macrolide resistance were unavailable. Therefore, we included the remaining 16 patients (Figure 1).

**Figure 1.**
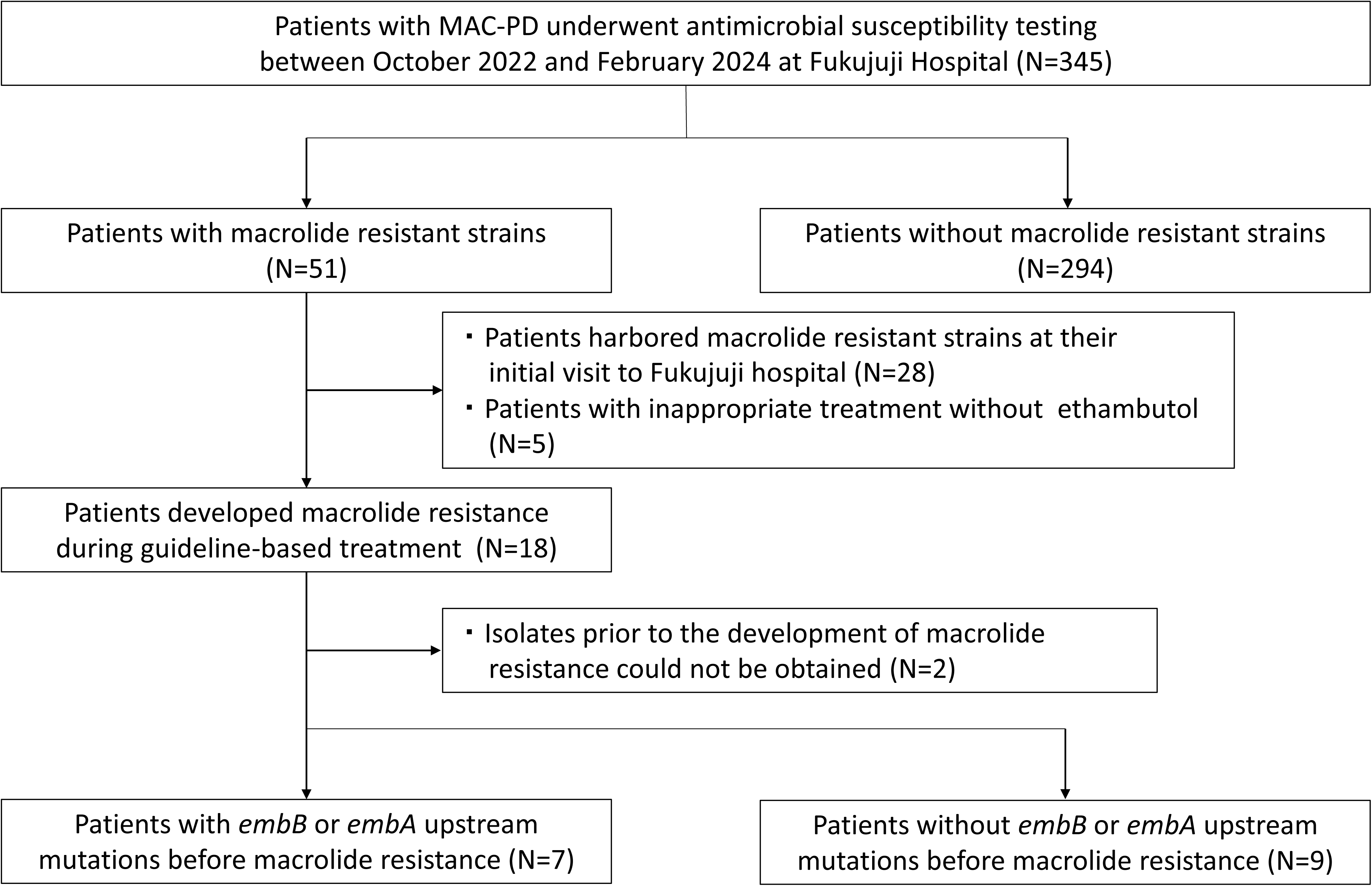
Flow diagram of the patients in this study. MAC-PD, *Mycobacterium avium* complex pulmonary disease

At macrolide resistance detection, we collected patient characteristics, acid-fast bacilli sputum culture and smear results, percent-predicted forced expiratory volume in 1 second (FEV1) and forced vital capacity (FVC), and chest computed tomography findings (cavities, radiological type, and severity). In addition, treatment histories prior to the development of macrolide resistance were collected.

Furthermore, for each patient, isolates obtained before treatment initiation, prior to the development of macrolide resistance, at the time of macrolide resistance detection, and after the development of macrolide resistance were collected. For each isolate, the MICs of clarithromycin, ethambutol, and rifampicin were determined, and genetic analyses were performed to detect *rrl* mutations associated with clarithromycin resistance and *embB* or *embA* upstream mutations. We examined whether *embB* or *embA* upstream mutations preceded the development of macrolide resistance and compared clinical, microbiological, and radiological data between patients with and without these mutations.

The institutional review board of Fukujuji Hospital (#25025) gave ethical approval for this work. Informed consent was obtained using an opt-out approach.

### Radiological evaluations

A specialized respiratory physician evaluated the most recent chest computed tomography findings obtained prior to the detection of macrolide resistance. Radiographic patterns were classified as noncavitary nodular bronchiectatic type, cavitary nodular bronchiectatic type, fibrocavitary type, or unclassifiable [14]. Radiological severity was assessed using the NICE scoring system, a visual scoring method for NTM-PD [15].

### Antimicrobial susceptibility testing

Antimicrobial susceptibility testing was conducted using the broth microdilution method recommended by Clinical and Laboratory Standards Institute (CLSI) guidelines, with cation-adjusted Mueller–Hinton broth (CAMHB) supplemented with oleic acid–albumin–dextrose–catalase (OADC) [16]. MICs were interpreted after the samples were incubated at 36 ± 1 °C in room air for 7 days. The tested concentrations ranged from 0.25 to 64 μg/mL for clarithromycin, 2 to 64 μg/mL for azithromycin, 2 to 128 μg/mL for ethambutol, and 0.25 to 8 μg/mL for rifampicin. Macrolide resistance was defined as resistance to clarithromycin, with clarithromycin resistance defined as an MIC ≥32 μg/mL [16].

### Genetic analysis

DNA was extracted as described previously [17]. Libraries were prepared using the QIAseq FX DNA library kit (Qiagen, Venlo, Netherlands), and 150-bp paired-end sequencing was performed on the NextSeq 550 System (Illumina, California, USA). Genomic analysis was performed as described previously [18]. Briefly, the sequence reads were aligned to OCU901 (RefSeq accession number GCF_002716925.3), and single-nucleotide polymorphisms were identified for each patient.

We analysed SNP distances, and isolates with ≤25 SNPs were regarded as the same strain. SNP analysis revealed that, in 2 of the 16 cases (FCR 4 and FCR 16), the isolates obtained before treatment initiation were genetically distinct from those obtained during treatment for MAC-PD. In addition, in one case (FCR 10), the isolate obtained after the detection of macrolide resistance was distinct from those obtained before and at the time of macrolide resistance detection. These isolates were excluded from the longitudinal analysis.

### Time-kill assay

To test the hypothesis that isolates acquiring ethambutol resistance mutations after treatment initiation—while remaining macrolide susceptible—are more prone to develop macrolide resistance than pretreatment isolates, we performed *in vitro* time-kill assays using recently reported methods [13]. In brief, bottles containing Middlebrook 7H9 broth with 0.05% Tween 80 and 10% OADC were inoculated with *M. avium* isolates. Azithromycin was added at a concentration of 4× MIC, which is the condition necessary to reproducibly develop stable macrolide resistance due to point mutations in the *rrl* gene. Under these conditions, ethambutol was tested at concentrations ranging from 0.03× to 2× the MIC to assess the suppression of macrolide resistance emergence. All experiments were performed in quadruplicate at 37 °C with shaking at 135 rpm. The numbers of total bacteria and macrolide-resistant bacteria were quantified using Middlebrook 7H10 agar plates and 32 mg/L clarithromycin-containing plates, respectively, in accordance with the CLSI criteria [16]. The experiments were conducted using paired isolates consisting of pretreatment strains and corresponding isolates that acquired ethambutol resistance solely because of *embB* mutations during treatment, while remaining macrolide susceptible.

### Statistical analysis

All data are expressed as numbers (percentages) for categorical variables and as medians [interquartile ranges (IQRs)] for continuous variables. Fisher’s exact test was applied to categorical variables, and the Mann–Whitney U test was applied to continuous variables. Clinical, microbiological, and radiological data were compared between patients with and without *embB* or *embA* upstream mutations prior to the detection of macrolide resistance. All statistical analyses were performed using EZR version 1.68 (Saitama Medical Center, Jichi Medical University, Saitama, Japan) [19].

## Results

In the 16 patients who developed macrolide resistance during guideline-based treatment, the median time from the initiation of MAC-PD treatment to the detection of macrolide resistance was 55 months (IQR, 39–60 months). The clinical characteristics at the time of macrolide resistance detection were as follows: median age, 65.5 years (IQR, 60.3–78.5 years); female, 14/17 (87.5%); median body mass index (BMI), 17.8 kg/m² (IQR, 16.3–19.2 kg/m²); history of smoking, 4/16 (25.0%); *M. avium*, 15/16 (93.8%); and presence of cavities, 10/16 (62.5%) (Table 1).

**Table 1.** Patient characteristics at the time of macrolide resistance detection. BMI, body mass index; FVC, forced vital capacity; FEV_1_, forced expiratory volume in 1 second.

|  | Total, N=16 | With *embB* or *embA*<br>upstream mutations<br>before macrolide<br>resistance, N=7 | Without *embB* or *embA*<br>upstream mutations<br>before macrolide<br>resistance, N=9 | P value |
| --- | --- | --- | --- | --- |
| Age, median [IQR]<br>(years) | 65.5 [60.3, 78.5] | 64.0 [60.5, 72.0] | 68.0 [61.0, 80.0] | 0.672 |
| Female, n (%) | 14 [87.5] | 7 (100) | 7 (77.8) | 0.475 |
| BMI, median [IQR]<br>(kg/m<sup>2</sup>) | 17.8 [16.3, 19.2] | 17.9 [17.7, 18.3] | 16.4 [15.0, 20.0] | 0.376 |
| Smoking history, n<br>(%) | 4 (25.0) | 2 (28.6) | 2 (22.2) | 1.000 |
| *M. avium*, n (%) | 15 (93.8) | 7 (100) | 8 (88.9) | 1.000 |
| Sputum smear<br>positive, n (%) | 8 (50.0) | 3 (42.9) | 5 (55.6) | 1.000 |
| Percent predicted<br>FVC, median [IQR]<br>(%) | 82.8 [63.9, 95.5] | 95.1 [94.0, 109.2] | 66.7 [63.1, 89.5] | 0.147 |
| Percent predicted<br>FEV<sub>1</sub>, median [IQR]<br>(%) | 79.7 [55.7, 93.3] | 112.1 [90.5, 114.5] | 78.5 [49.4, 79.9] | 0.059 |
| Cavities, n (%) | 10 (62.5) | 1 (14.3) | 9 (100) | <0.001 |
| NICE score, median<br>[IQR] | 14[8, 19] | 12 [7, 19] | 14 [8, 18] | 0.560 |
| Underlying disease: |  |  |  |  |
| Sinusitis, n (%) | 2 (12.5) | 0 (0) | 2 (22.2) | 0.475 |
| Diabetes mellitus, n<br>(%) | 3 (18.8) | 0 (0) | 3 (33.3) | 0.213 |
| Gastroesophageal<br>reflux disease, n (%) | 3 (18.8) | 0 (0) | 3 (33.3) | 0.213 |
| Post tuberculosis, n<br>(%) | 1 (6.3) | 1 (14.3) | 0 (0) | 0.438 |
| Lung cancer, n (%) | 1 (6.3) | 0 (0) | 1 (11.1) | 1.000 |
| Treatment duration<br>before macrolide<br>resistance, median<br>[IQR] (months) | 55 [39, 60] | 52 [38, 56] | 60 [41, 62] | 0.168 |

### Longitudinal changes in MICs

Although none of the patients had an ethambutol MIC ≥16 μg/mL before treatment initiation, 8 had an ethambutol MIC ≥16 μg/mL at the time macrolide resistance was identified (Supplementary Table S1; Figure 2A). Of these patients, seven already exhibited an ethambutol MIC ≥16 μg/mL prior to the development of macrolide resistance (Table 2).

**Figure 2.**
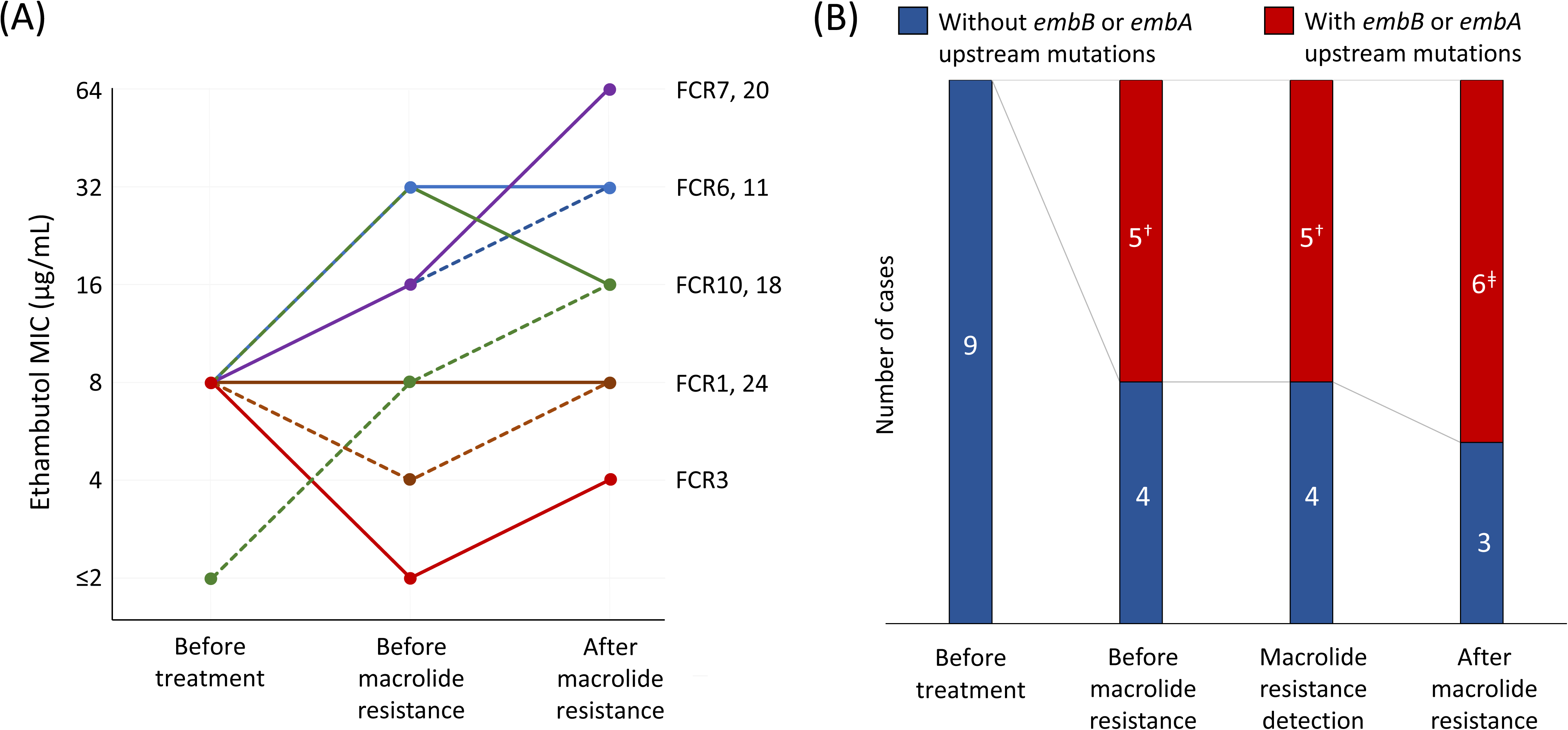
(A) Longitudinal changes in the minimum inhibitory concentration of ethambutol. (B) Frequencies of *embB* or *embA* upstream mutations at each time point. The numbers in the bars indicate the number of cases. * Cases with an ethambutol MIC less than 2 μg/mL were also included. †In one case, an *embB* mutation was detected before macrolide resistance, whereas an *embA* upstream mutation was observed in isolates at the time of the development of macrolide resistance and after macrolide resistance. ‡In one case, the *embB* mutation was first detected in an isolate obtained after the development of macrolide resistance. MIC, minimum inhibitory concentration

**Table 2.** Ethambutol MICs and *embB* or *embA* upstream mutations before and after the development of macrolide resistance. MIC, minimum inhibitory concentration.

|  |  | Before macrolide resistance | After development of<br>macrolide resistance |
| --- | --- | --- | --- |
| FCR-2 | Ethambutol, MIC (µg/mL) | 16 | 16 |
|  | *embB* or *embA* upstream<br>mutations | *embA* upstream | - |
| FCR-6 | Ethambutol, MIC (µg/mL) | 16 | 32 |
|  | *embB* or *embA* upstream<br>mutations | *embB* | *embB* |
| FCR-7 | Ethambutol, MIC (µg/mL) | 16 | 64 |
|  | *embB* or *embA* upstream<br>mutations | *embB* | *embB/ embA* upstream |
| FCR-8 | Ethambutol, MIC (µg/mL) | 16 | 32 |
|  | *embB* or *embA* upstream<br>mutations | *embB* | *embB* |
| FCR-10 | Ethambutol, MIC (µg/mL) | 32 | 16 |
|  | *embB* or *embA* upstream<br>mutations | *embB* | *embB* |
| FCR-11 | Ethambutol, MIC (µg/mL) | 32 | 32 |
|  | *embB* or *embA* upstream<br>mutations | *embB* | *embB* |
| FCR-20 | Ethambutol, MIC (µg/mL) | 16 | 64 |
|  | *embB* or *embA* upstream<br>mutations | *embB/ embA* upstream | *embB/ embA* upstream |
| FCR-18 | Ethambutol, MIC (µg/mL) | 8 | 16 |
|  | *embB* or *embA* upstream<br>mutations | - | - |
| FCR-1 | Ethambutol, MIC (µg/mL) | 8 | 4 |
|  | *embB* or *embA* upstream<br>mutations | - | - |
| FCR-3 | Ethambutol, MIC (µg/mL) | ≤2 | 4 |
|  | *embB* or *embA* upstream mutations | - | - |
| FCR-4 | Ethambutol, MIC (µg/mL) | ≤2 | 8 |
|  | *embB* or *embA* upstream mutations | - | - |
| FCR-5 | Ethambutol, MIC (µg/mL) | 4 | 4 |
|  | *embB* or *embA* upstream mutations | - | - |
| FCR-9 | Ethambutol, MIC (µg/mL) | 8 | 4 |
|  | *embB* or *embA* upstream mutations | - | - |
| FCR-16 | Ethambutol, MIC (µg/mL) | 8 | 8 |
|  | *embB* or *embA* upstream mutations | - | - |
| FCR-24 | Ethambutol, MIC (µg/mL) | 4 | 8 |
|  | *embB* or *embA* upstream mutations | - | - |
| FCR-25 | Ethambutol, MIC (µg/mL) | ≤2 | ≤2 |
|  | *embB* or *embA* upstream mutations | - | - |

Ethambutol MICs tended to increase throughout the course of treatment (Figure 2A; Supplementary Table S1). In contrast, rifampicin MICs did not increase throughout the course of treatment (Supplementary Table S1; Supplementary Figure S1).

### Longitudinal changes in embB or embA upstream mutations

Although no isolates harbored *embB* or *embA* upstream mutations before treatment initiation, 7/16 (43.8%) had *embB* or *embA* upstream mutations prior to the development of macrolide resistance (Table 1). Longitudinal changes in *embB* or *embA* upstream mutations were also analysed in the nine patients for whom MIC trends were available. Five patients acquired *embB* or *embA* upstream mutations prior to the development of macrolide resistance, whereas one patient (FCR-24) acquired an *embB* mutation after the development of macrolide resistance (Supplementary Table S1; Figure 2B).

The median intervals from treatment initiation to the detection of *embB* or *embA* upstream mutations and from their detection to the development of macrolide resistance were 27 months (IQR, 23–34 months) and 16 months (IQR, 12–24 months), respectively.

### Comparison of clinical data between patients with and without preceding ethambutol resistance

Patients with *embB* or *embA* upstream mutations prior to the development of macrolide resistance exhibited a significantly higher rate of noncavitary nodular bronchiectatic type (85.7% vs. 0%, p < 0.001) and a significantly lower rate of cavitary lesions (14.3% vs. 100%, p < 0.001) than those without such mutations (Table 1). Treatment regimens and drug dosages were similar between patients with and without *embB* or *embA* upstream mutations (Table 3; Supplementary Figure S2).

**Table 3.**
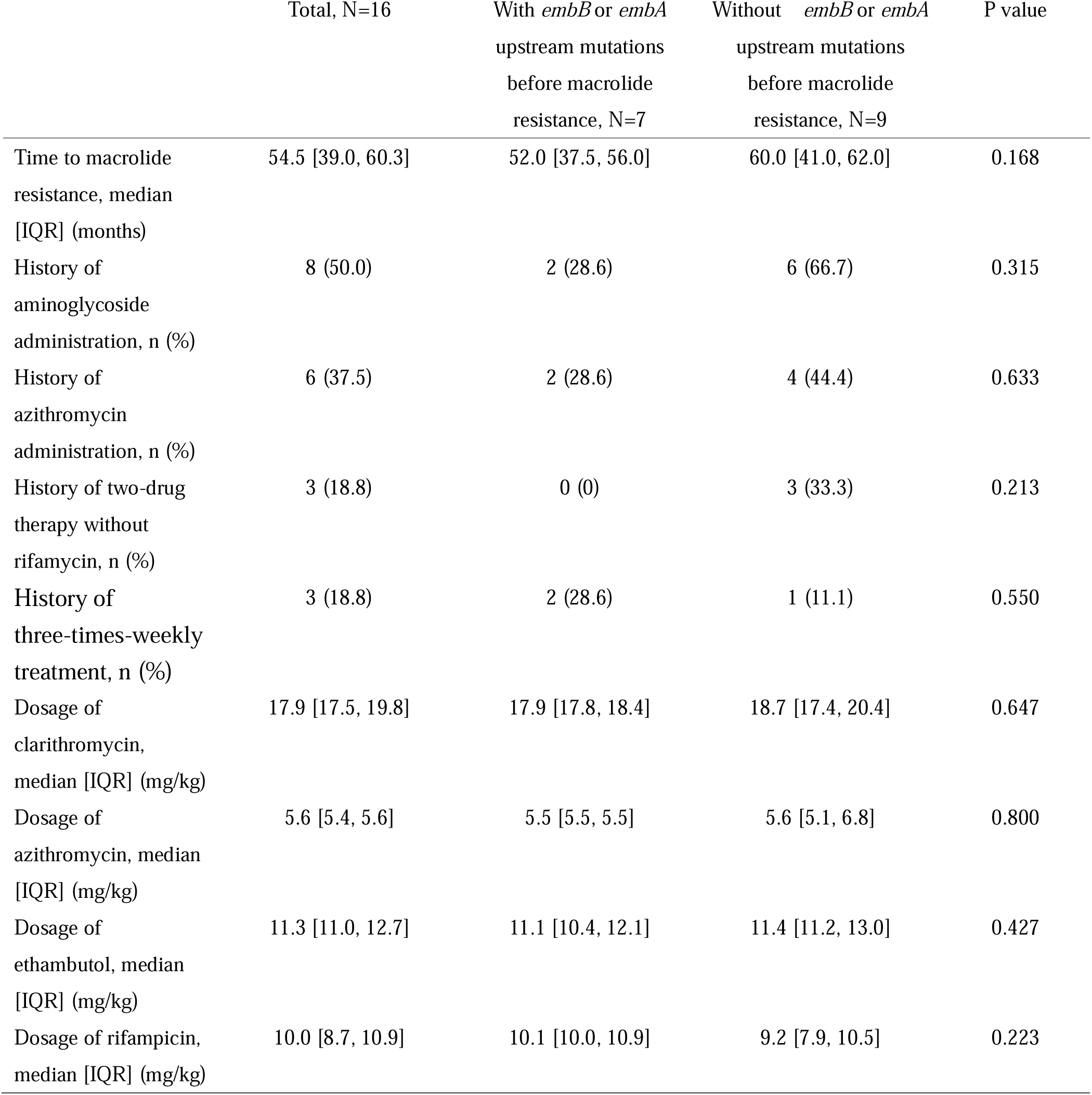
Treatment courses and drug dosages stratified by the presence or absence of *embB* or the upstream region of *embA* mutations prior to macrolide resistance.

### Time-kill assays

Among the five paired pretreatment and *embB*-mutant isolates, two pairs were excluded because of poor growth, and the remaining six strains from three pretreatment and *embB*-mutant pairs (FCR-6, -10, and -20) were included in the analysis. In strains with *embB* mutations, the ethambutol MIC increased by 1–2 dilutions relative to that of pretreatment isolates (Supplementary Table S1). The changes over time in the total and macrolide-resistant bacterial counts are presented in Supplementary Figure S3, and the relationships between ethambutol concentration and resistance counts at the final time point are shown in Figure 3. The emergence of macrolide resistance progressively decreased with increasing ethambutol concentration. In all isolate pairs, compared with the corresponding pretreatment strains, the *embB*-mutant strains required higher ethambutol concentrations to suppress macrolide resistance. Notably, complete suppression of macrolide resistance was achieved at 0.25–0.5× the ethambutol MIC in pretreatment isolates, whereas concentrations of 1–2× the MIC were required in *embB*-mutant isolates (Figure 3).

**Figure 3.**
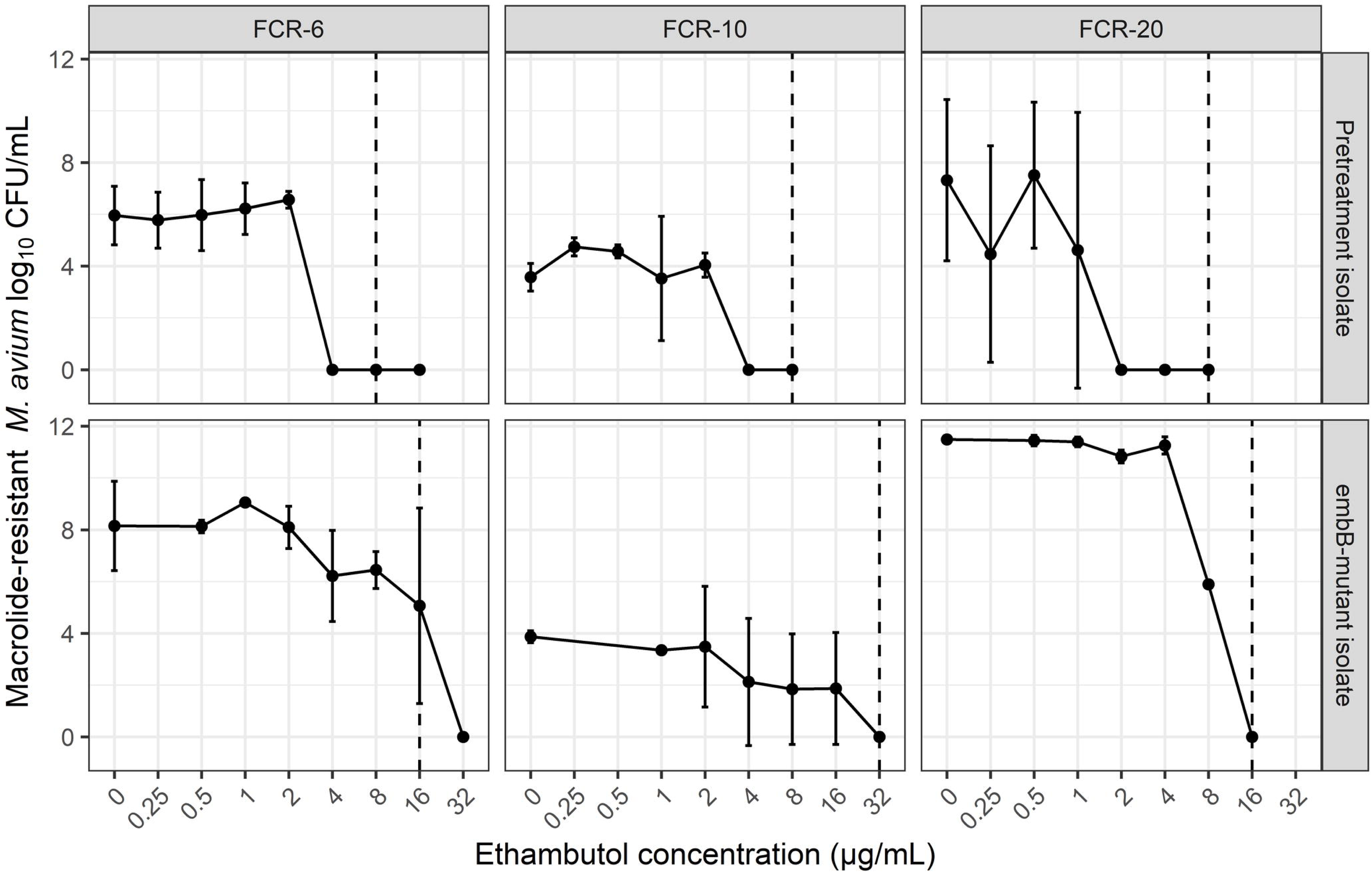
Macrolide-resistant bacterial counts after 28 days of culture in time-kill assays. The solid points and lines represent the mean values, and the error bars indicate the standard deviations. Dashed lines indicate the MIC for each isolate.

## Discussion

To the best of our knowledge, this is the first report to demonstrate cases in which the acquisition of mutations associated with ethambutol resistance preceded the development of macrolide resistance during guideline-based treatment for MAC-PD. Although no patients had an ethambutol MIC ≥16 μg/mL or *embB* or *embA* upstream mutations at treatment initiation, 55.6% showed an increase in the ethambutol MIC to ≥16 μg/mL, and 43.8% acquired *embB* or *embA* upstream mutations prior to the development of macrolide resistance. Patients who developed *embB* or *embA* upstream mutations preceding macrolide resistance exhibited a significantly lower frequency of cavitary lesions than those without such mutations (14.3% vs. 100%, p < 0.001).

The impact of ethambutol MICs on treatment outcomes in patients with MAC-PD remains controversial. Previous studies have reported that ethambutol MICs ≥8 μg/mL are associated with lower culture conversion and treatment success rates [20, 21]. In contrast, Moon et al. demonstrated that even when the MICs of ethambutol were ≥8 μg/mL, culture conversion and microbiological cure rates were not significantly affected [22]. In the present study, *embB* or *embA* upstream mutations were detected in only 1 of 11 isolates (9.1%) with an ethambutol MIC of 8 μg/mL but in 12 of 15 isolates (80%) with an ethambutol MIC ≥16 μg/mL (Supplementary Table S1). These findings suggest that an ethambutol MIC cutoff of 16 μg/mL may provide a more appropriate criterion for evaluating the impact of ethambutol resistance on clinical outcomes in patients with MAC-PD.

Cavitary lesions and acid-fast bacilli smear positivity have been reported to be risk factors for the development of macrolide resistance [9, 10]. However, in the present study, 37.5% of patients had no cavitary lesions, and 50.0% were acid-fast bacilli smear-negative at the time macrolide resistance developed. Notably, *embB* or *embA* upstream mutations preceding macrolide resistance were identified in all noncavitary patients with MAC-PD (6/6, 100%). These findings suggest that prior ethambutol resistance may partly explain the development of macrolide resistance in patients without cavitary lesions.

On the other hand, in patients with cavities, *embB* or *embA* upstream mutations were detected in only 10% of cases. These findings suggest that MAC-PD patients with cavities, mechanisms other than the acquisition of *embB* or *embA* upstream mutations are likely to contribute to the development of macrolide resistance. In addition to the selection of resistant strains due to a high mycobacterial burden, poor drug penetration into cavities may also contribute to macrolide resistance by creating a situation similar to that of clarithromycin monotherapy due to inadequate ethambutol penetration. However, most prior studies of drug distribution were conducted not in NTM models but in tuberculosis patients or animal models [23, 24]. Although the distribution of macrolides into cavities has been studied using lung tissue resected from patients with NTM-PD [25], the distribution of ethambutol in patients with NTM-PD has not yet been elucidated.

In the time-kill experiments, increases in MICs of ethambutol associated with *embB* mutations were accompanied by a requirement for higher ethambutol concentrations to suppress the emergence of macrolide resistance. Complete suppression was achieved at 0.25–0.5× the MIC in pretreatment isolates, whereas concentrations of 1–2× the MIC were required in *embB*-mutant isolates, suggesting that *embB*-mutant strains may require ethambutol exposures exceeding those predicted solely by MIC elevation. Our findings suggest that *embB* or *embA* upstream mutations markedly increase the ethambutol concentrations required for resistance suppression, indicating that additional companion drugs, such as amikacin liposome inhalation suspension or parenteral aminoglycosides, may be required in such cases.

This study had several limitations. First, the number of patients who developed macrolide resistance during guideline-based treatment and underwent genetic analysis was relatively small. However, considering that the frequency of macrolide resistance during guideline-based treatment has been reported to be 9–12% [9–11], the number of eligible patients in this study is reasonable for a single-center analysis. Second, although we demonstrated cases in which mutations associated with ethambutol resistance preceded macrolide resistance, this study could not confirm whether such mutations themselves increase the risk of macrolide resistance. Further basic research and prospective clinical studies are needed to validate this relationship.

## Conclusion

In conclusion, the acquisition of *embB* or *embA* upstream mutations may contribute to the development of macrolide resistance, particularly in patients without cavities. Regular monitoring of ethambutol MICs, especially MICs of ≥16 µg/mL, and *embB* or *embA* upstream mutations may help predict the development of macrolide resistance. Furthermore, establishing effective treatment strategies both to prevent *embB* or *embA* upstream mutations and to manage patients with these mutations may help reduce the risk of macrolide resistance in refractory MAC-PD.

## CRediT authorship contribution statement

Masashi Ito: Conceptualization, Methodology, Investigation, Data curation, Formal analysis, Writing – original draft. Fumiya Watanabe: Investigation, Writing – original draft. Asami Osugi: Investigation, Writing – review & editing. Akio Aono: Investigation, Writing – review & editing. Keiji Fujiwara: Writing – review & editing. Koji Furuuchi: Writing – review & editing. Tatsuya Kodama: Writing – review & editing. Takashi Ohe: Writing – review & editing. Takashi Yoshiyama: Writing – review & editing. Shoji Kudoh: Writing – review & editing. Satoshi Mitarai: Methodology, Writing – review & editing. Kozo Morimoto: Conceptualization, Methodology, Investigation, Supervision, Writing – review & editing. All authors approved the final version of the manuscript.

## Acknowledgements

We thank Kazue Mizuno and Akira Ito for their assistance with the bacteriological analyses.

## Data availability statement

The sequence data generated in this study have been deposited in the NCBI BioProject database under accession number PRJNA1468305.

