## Supplemental files for "Ethambutol resistance preceding macrolide resistance in *Mycobacterium avium* complex pulmonary disease: a retrospective longitudinal study and in vitro analysis"

**Supplementary Table**

**Table S1:** Longitudinal changes in the MICs of clarithromycin, ethambutol, and rifampicin, and mutations in the *embB*, the upstream region of *embA*, and *rrl* genes. Sections with an ethambutol MIC ≥16 μg/mL or with detected mutations in *embB* or *embA* are shaded in gray and emphasized in bold.

MIC, minimum inhibitory concentration; NE, not evaluated

* The *embB Tyr288Ser* mutation detected in FCR-10 has not been reported to be associated with ethambutol resistance, and its contribution to resistance remains uncertain.

| Cases | | Before Treatment | Before macrolide resistance | Macrolide resistance detection | After macrolide resistance |
|---|---|---|---|---|---|
| FCR-1 | Clarithromycin, MIC (µg/mL) | 1 | 1 | >64 | >64 |
| | *rrl* mutation | - | - | NE | 2059 |
| | Ethambutol, MIC (µg/mL) | 8 | 8 | 8 | 4 |
| | *embB* or *embA* upstream mutation | - | - | NE | - |
| | Rifampicin, MIC (µg/mL) | 8 | 2 | 1 | 4 |
| FCR-3 | Clarithromycin, MIC (µg/mL) | 0.5 | 1 | >64 | >64 |
| | *rrl* mutation | - | - | 2059 | 2059 |
| | Ethambutol, MIC (µg/mL) | 8 | ≤2 | 4 | 4 |
| | *embB* or *embA* upstream mutation | - | - | - | - |
| | Rifampicin, MIC (µg/mL) | 2 | 2 | 4 | 4 |
| FCR-6 | Clarithromycin, MIC (µg/mL) | 0.5 | 1 | >64 | >64 |
| | Azithromycin, MIC (µg/mL) | 8 | 8 | NE | NE |
| | *rrl* mutation | - | - | 2059 | 2059 |
| | Ethambutol, MIC (µg/mL) | 8 | 16 | 32 | 32 |
| | *embB* or *embA* upstream mutation | - | *embB_Gln497Arg* | *embB_Gln497Arg* | *embB_Gln497Arg* |
| | Rifampicin, MIC (µg/mL) | 1 | 0.5 | 0.5 | 0.5 |
| FCR-7 | Clarithromycin, MIC (µg/mL) | 4 | 2 | >64 | >64 |
| | *rrl* mutation | - | - | 2058 | 2058 |
| | Ethambutol, MIC (µg/mL) | 8 | 16 | 64 | 32 |
| | *embB* or *embA* upstream mutation | - | *embB_Met306Thr* | *embB_Met306Thr/embA_-12* | *embA_-12* |
| | Rifampicin, MIC (µg/mL) | >8 | >8 | 4 | 2 |

| | | | | | |
|---|---|---|---|---|---|
| FCR-10 | Clarithromycin, MIC (µg/mL) | 1 | 0.5 | >64 | NE |
| | Azithromycin, MIC (µg/mL) | 64 | 64 | NE | NE |
| | *rrl* mutation | - | - | 2058/2059 | NE |
| | Ethambutol, MIC (µg/mL) | 8 | 32 | 16 | NE |
| | *embB* or *embA* upstream mutation | - | *embB_Tyr288 Ser* | *embB_ Tyr288Ser** | NE |
| | Rifampicin, MIC (µg/mL) | 1 | 2 | 1 | NE |
| FCR-11 | Clarithromycin, MIC (µg/mL) | 1 | 2 | >64 | >64 |
| | *rrl* mutation | - | - | 2059 | 2059 |
| | Ethambutol, MIC (µg/mL) | 8 | **32** | **32** | **32** |
| | *embB* or *embA* upstream mutation | - | - | - | - |
| | Rifampicin, MIC (µg/mL) | 4 | >8 | 1 | >8 |
| FCR-18 | Clarithromycin, MIC (µg/mL) | 0.5 | 0.25 | >64 | NE |
| | *rrl* mutation | - | - | 2058 | NE |
| | Ethambutol, MIC (µg/mL) | ≤2 | 8 | 16 | NE |
| | *embB* or *embA* upstream mutation | - | *embB_ Met306Ile* | *embB_ Met306Ile* | NE |
| | Rifampicin, MIC (µg/mL) | 0.5 | 0.25 | 0.5 | NE |
| FCR-20 | Clarithromycin, MIC (µg/mL) | 1 | 0.5 | >64 | >64 |
| | Azithromycin, MIC (µg/mL) | 4 | 8 | | |
| | *rrl* mutation | - | - | - | 2058 |
| | Ethambutol, MIC (µg/mL) | 8 | 16 | 64 | 32 |
| | *embB* or *embA* upstream mutation | - | *embB_ Gln497Arg/ embA_-12* | *embB_ Gln497Arg/ embA_-12* | *embB_ Gln497Arg/ embA_-12* |
| | Rifampicin, MIC (µg/mL) | 4 | 0.5 | 8 | 2 |

| | | | | | |
|---|---|---|---|---|---|
| FCR-24 | Clarithromycin, MIC (µg/mL) | 0.5 | 0.5 | >64 | >64 |
| | *rrl* mutation | - | - | 2058 | 2058 |
| | Ethambutol, MIC (µg/mL) | 8 | 4 | 8 | **32** |
| | *embB* or *embA* upstream mutation | - | - | - | *embB_ Met306Ile* |
| | Rifampicin, MIC (µg/mL) | 1 | 1 | 0.5 | 1 |

**Supplementary Figures**

**Figure S1:** Longitudinal changes in the MICs of rifampicin from pretreatment to the development of macrolide resistance.

*In FCR-7, the MIC of rifampicin was >8 μg/mL in isolates obtained both before treatment initiation and prior to the development of clarithromycin resistance. In FCR-11, the MIC of rifampicin in isolates obtained prior to the development of clarithromycin resistance was also >8 μg/mL.

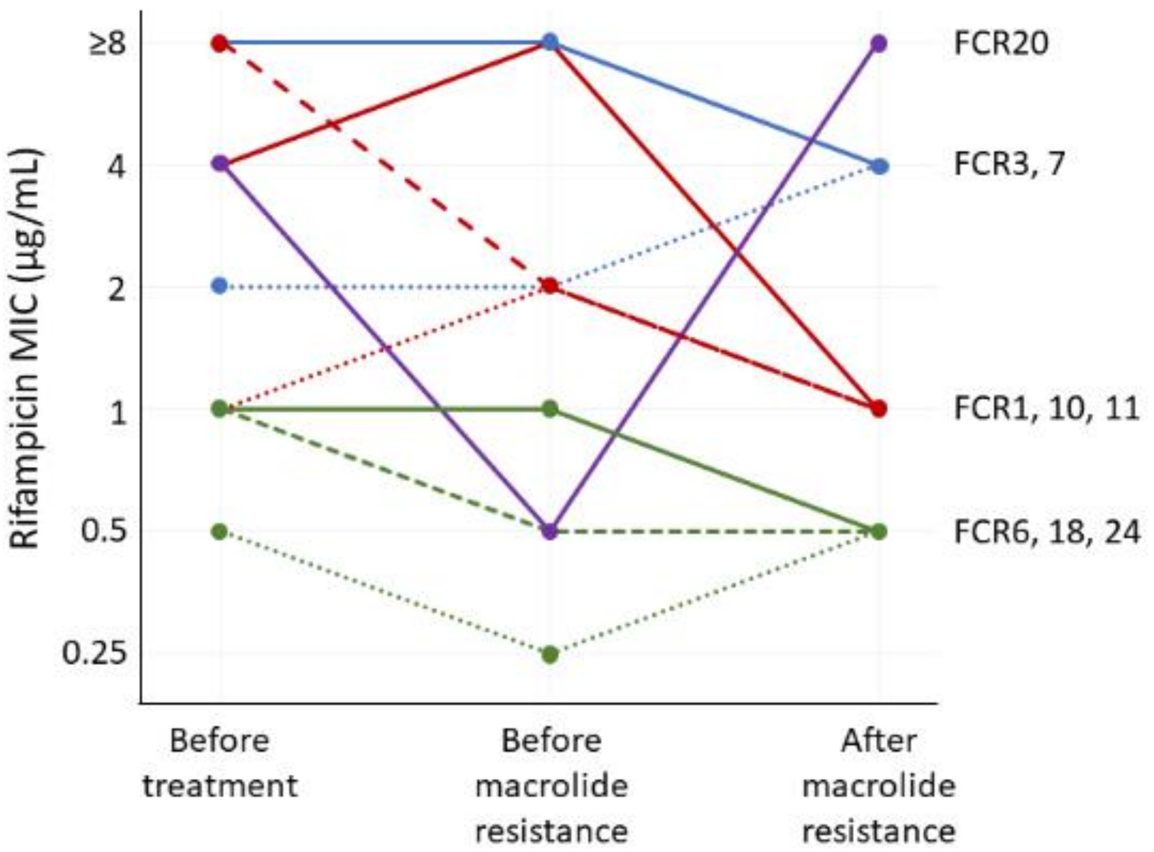

**Figure S2:** Treatment courses of each patient from the initiation of treatment to the detection of macrolide resistance. Black arrows indicate the time points at which isolates were obtained prior to the development of macrolide resistance for drug susceptibility testing and genetic analysis. The numbers shown in parentheses after each regimen indicate the treatment duration of that regimen.

C, clarithromycin; E, ethambutol; R, rifampicin; A, azithromycin; AI, amikacin infusion; TIW, three-times-weekly; AL, amikacin liposome inhalation suspension; CL, clofazimine; ST, sitafloxacin

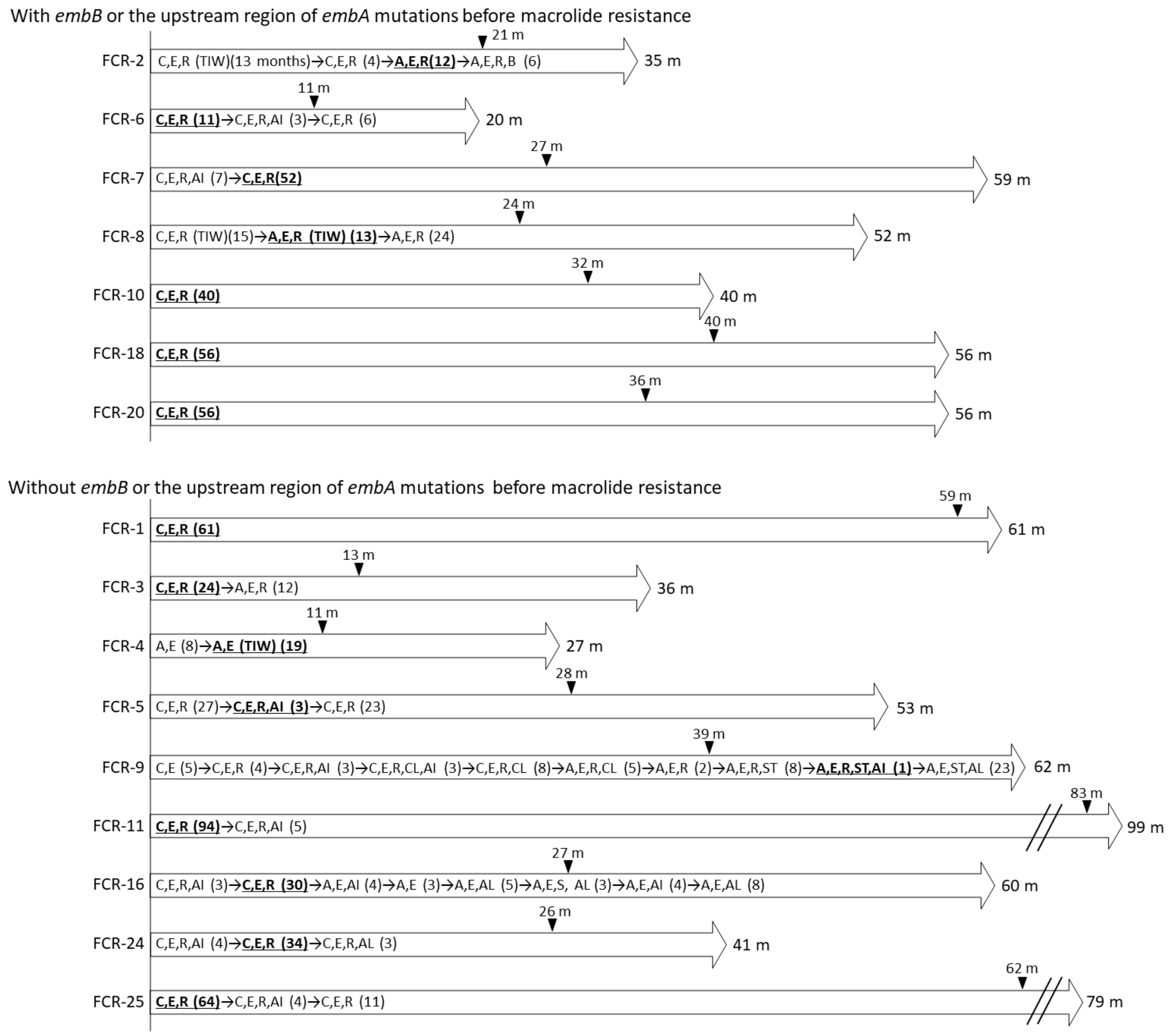

**Figure S3:** Changes in the number of total and macrolide-resistant bacteria.

6-1 and 6-2 represented pretreatment and *embB* mutant strain of FCR-6, respectively. 10-1 and 10-2 represented pretreatment and *embB* mutant strain of FCR-10, respectively. 20-1 and 20-2 represented pretreatment and *embB* mutant strain of FCR-20, respectively. Y-axis represents the logarithmic *M. avium* concentration and X-axis indicates the time elapsed after the start of incubation. Each experiment was performed in quadruplicate. Grey dots and lines represent the total bacterial counts, whereas black dots and lines represent the counts of resistant bacteria. All conditions included azithromycin at 4× MIC and the column concentrations represented co-incubated ethambutol concentrations.

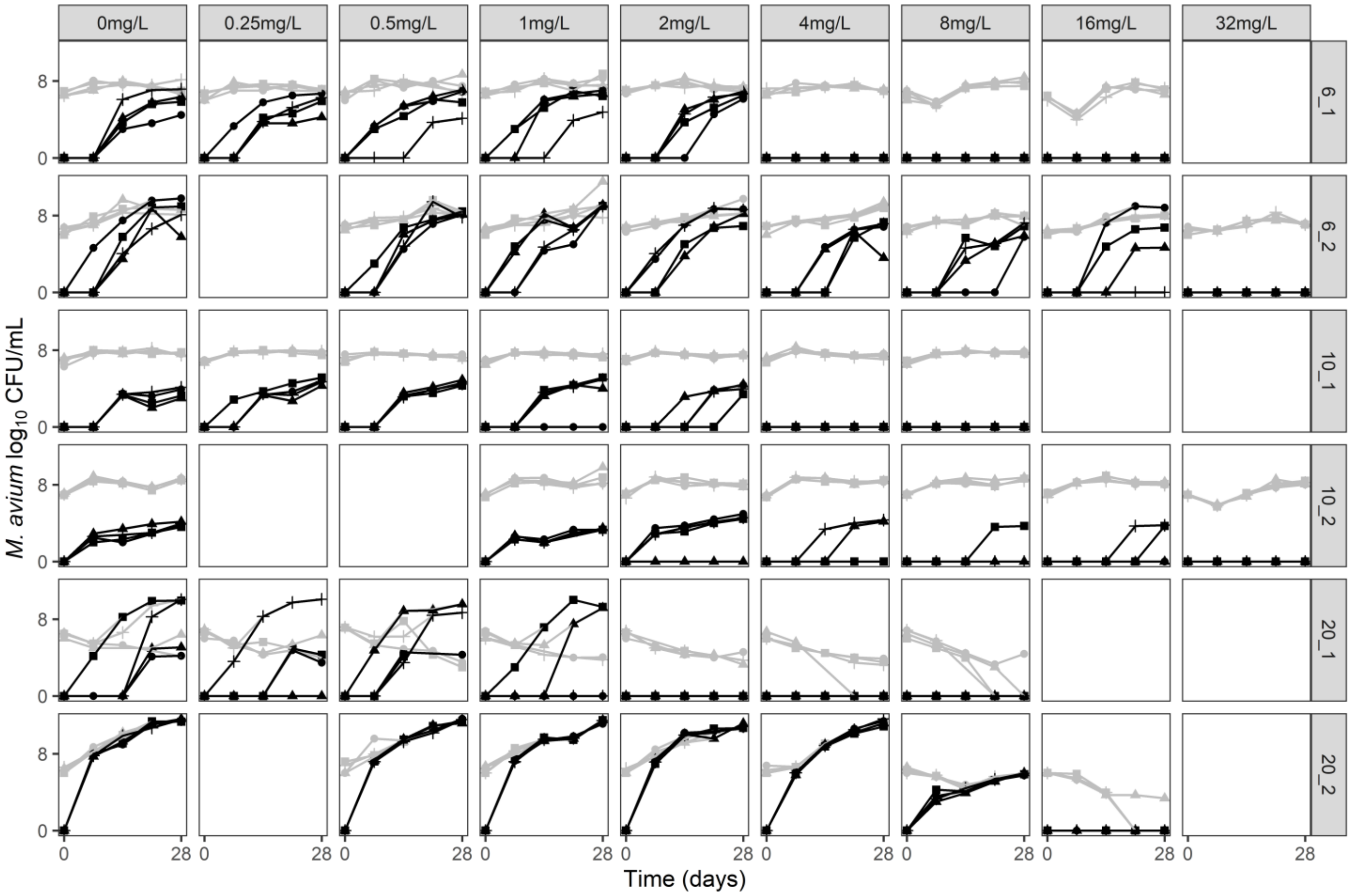
